# Evidence-based public health messaging on the non-visual effects of ocular light exposure: A modified Delphi expert consensus

**DOI:** 10.1101/2025.05.07.25327160

**Authors:** Manuel Spitschan, Laura Kervezee, Oliver Stefani, Marijke Gordijn, Jennifer A. Veitch, Renske Lok, the Light for Public Health Consortium

## Abstract

**Introduction:** In addition to vision, light regulates circadian rhythms, sleep, mood, and alertness. Despite growing scientific understanding, there remains a gap in translating this knowledge into accessible, evidence-based guidance. The goal of this paper is to formulate scientifically grounded statements about the influence of light on human psychological and physiological health, intended for dissemination to the public and policymakers.

**Methods:** An international consortium of 21 experts convened at the Ladenburg Roundtable in April 2024. Experts were selected based on their scientific contributions to chronobiology, psychology, neuroscience, and the measurement and practical application of light. Through a structured, iterative modified Delphi process, 27 statements were developed. Each statement included a simplified public-facing version and contextual information to support understanding. Consensus was assessed using predefined thresholds of agreement (>75% endorsement). Statements not meeting consensus were revised and re-evaluated.

**Results:** Of the 27 proposed statements, 26 reached the threshold for consensus, with high levels of agreement across diverse topics. One statement did not reach consensus due to insufficient scientific evidence and was excluded, while another was revised based on feedback and subsequently accepted. The iterative revision process significantly improved the clarity, accuracy, and accessibility of the final statements. A readability assessment showed an average sentence length of 14.8 words and a Flesch-Kincaid Grade Level of 8.6, indicating that the statements suit a broad, non-specialist audience. The final consensus statements are available at lightforpublichealth.org.

**Conclusions:** This expert consensus provides clear, accessible messages about how light affects human health. The statements offer a practical tool for public education and policymaking and can be used by public-health multipliers (e.g., schools, employers, healthcare providers, and urban planners) to promote healthier light exposure in daily life. They highlight the importance of recognizing light as a key factor in health, alongside sleep, nutrition, and physical activity.

**Key message:** *What is already known on this topic:* Light exposure has an impact on human health beyond vision, affecting sleep, alertness, and circadian rhythms. However, policy guidance on healthy lighting is limited.

*What this study adds:* This expert consensus document sets out 26 key messages on the health impacts of light in clear, accessible language.

*How this study might affect research, practice, or policy:* These messages provide policymakers with reliable guidance to help create healthy lighting environments. They can inform regulations, public awareness campaigns, and health promotion strategies.

## Introduction

Over the past 50 years, the scientific understanding of how light affects human health has advanced dramatically. Beyond enabling vision, light regulates core biological processes, including circadian rhythms, sleep, and hormone secretion. The discovery of intrinsically photosensitive retinal ganglion cells (ipRGCs) revealed how light exerts effects on human physiology and psychology^1–3^. These cells play a central role in synchronizing the biological clock to the external light-dark cycle^4,5^, thereby influencing sleep-wake timing^6,7^. Additionally, light acutely affects alertness^8,9^, melatonin production (e.g. ^9–11^), and cognitive performance^12,13^. These discoveries prompted the development of standards for measuring biologically effective light. In 2018, the International Commission on Illumination introduced CIE S 026/E:2018, providing a standardized framework for quantifying biologically effective ocular light^14^. In 2022, global experts released recommendations for healthy light exposure^15^. However, the translation of these findings into public health guidance remains a challenge, hindered by a lag in the adoption of research findings in policy and public health guidelines^16^.

To address this gap, 21 international experts convened at the Ladenburg Roundtable (April 14– 16, 2024, Germany) to develop consensus-based, evidence-informed statements on light and health. The goal was to support public health messaging, guide policy, inform architecture and workplace standards, and empower individuals. The resulting white paper identified seven key steps for effective communication, including simplifying technical language and aligning with existing public health recommendations^17^.

This report presents 26 consensus statements on the effects of light on human psychology and physiology, developed through an iterative review.

## Materials and Methods

### Ethics

This research involved expert participants contributing in their professional capacity to a consensus process. No clinical interventions, personal health data, or sensitive information were collected. Formal ethical approval was therefore not required under prevailing guidelines. All participants were informed about the purpose and procedures of the study and provided consent through their voluntary agreement to take part.

### Patient and Public Involvement statement

Patients or members of the public were not involved in the design, conduct, reporting, or dissemination of this research.

#### Participants

Participants for the Ladenburg Light & Health Roundtable were purposefully selected to ensure diversity in geography, career stage, and expertise. The final group included 25 experts from human clinical research, animal neuroscience, physiological optics, epidemiology, occupational health, environmental psychology, chronobiology, lighting science, and public health (Appendix A). They represented universities, research institutions, and public agencies. Four invited individuals were unable to attend. Of the 25 experts, 19 actively participated in the statement rating process. A coordinating committee of six lead authors representing five key organizations, the International Commission on Illumination (CIE), the Society for Light, Rhythms and Circadian Health (SLRCH), the Daylight Academy (DLA), the Good Light Group (GLG), and the Center for Environmental Therapeutics (CET), oversaw the process and endorsed the statements but did not contribute ratings. Participants were not compensated for their participation. All invited experts completed the process.

#### Statement development and consensus

We employed a modified Delphi technique involving two rounds of anonymized feedback using REDCap^18,19^ (Round 1) and via email-based commenting (Round 2), with a predefined consensus threshold of 75% agreement (**Figure 1)**. The development was not registered. The coordinating committee developed the initial statements, incorporating feedback from roundtable discussions. No systematic searches were conducted. In this development stage, the Committee members assessed the level of evidence using a three-alternative forced-choice procedure with three options (*low evidence*, i.e, based on preliminary or weak findings, limited studies, or inconsistent results; *moderate evidence* (i.e., supported by multiple studies but with some limitations, such as variability in results or study designs; *high evidence*, i.e., strongly supported by consistent, replicated research with large sample sizes and broad scientific consensus) Committee members simplified language, added contextual information, and cited relevant evidence. To minimize confirmation bias, however, all subsequent revisions were guided by anonymized ratings and open-text feedback from the broader group of expert participants, who were not members of the coordinating committee. This occurred through a two-step Delphi peer-review process: in Round 1, participants rated agreement (two-forced choice agree/disagree) and commented on the draft statements; feedback was integrated into revised versions. Participants reviewed simplified statements and contextual text in Round 2, ensuring scientific accuracy and clarity. The entire process, including consensus-building, peer review, and readability adjustments, occurred between October 2024 and February 2025.

**Figure 1.**
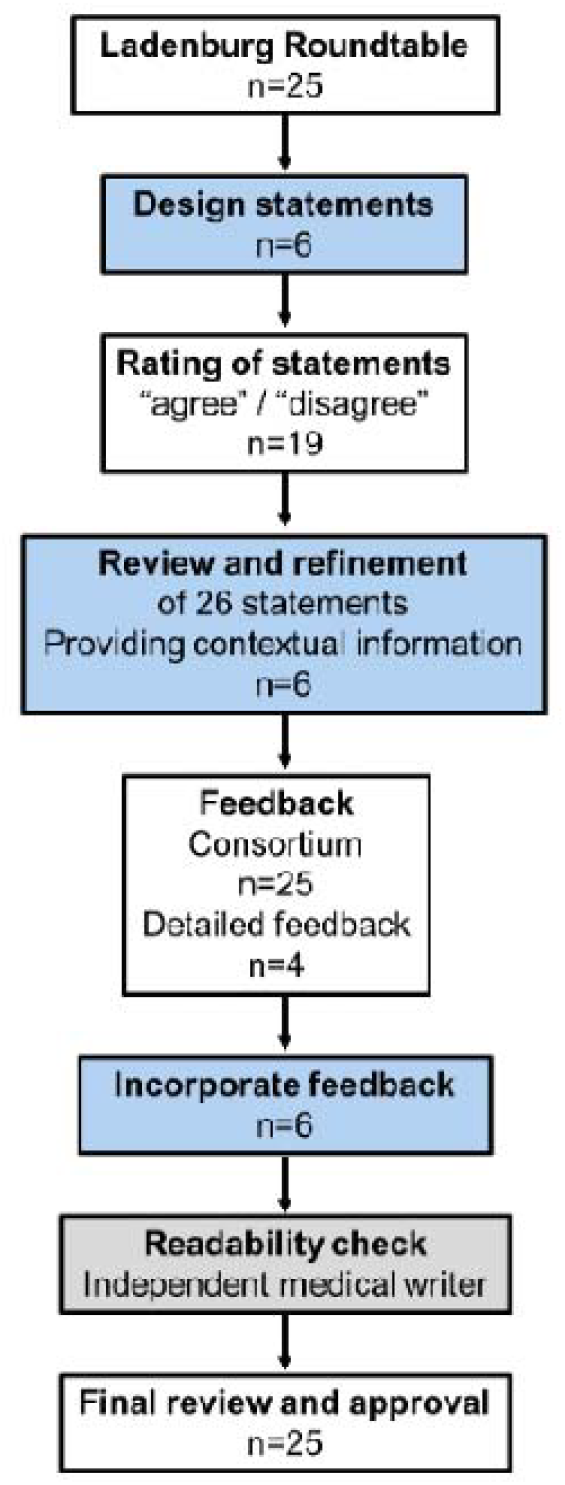
Flow chart. The coordinating committee led the initial development of key statements, which were then submitted for consensus approval by the Light for Public Health Consortium. Each statement received feedback, with in-depth input from four Consortium members incorporated into the revisions. Once the core statements were finalized, simplified versions and contextual information were developed, followed by an additional round of feedback. A medical writer then assessed the document for English readability, ensuring accessibility while preserving the original scientific statements. Finally, the Light for Public Health Consortium was given a final opportunity to review the scientific statements, along with their simplified versions and contextual details, before finalization. Boxes in light blue indicate steps led by the steering committee (n=6), white presents the Consortium (n=25) and grey the independent external medical writer (n=1).

#### Evidence rating

Each scientific statement developed in this consensus process was rated for evidence certainty following a structured approach. We based our assessment on a modified version of the Oxford Centre for Evidence-Based Medicine (OCEBM) Quality Rating Scheme for Studies and Other Evidence.

The certainty of evidence for each statement was assigned as follows:

- **Level 1 (High certainty)**: Properly powered and conducted randomized clinical trials, or systematic reviews with meta-analysis.
- **Level 2 (Moderate certainty)**: Well-designed controlled trials without randomization, or prospective comparative cohort studies.
- **Level 3 (Moderate to low certainty)**: Case-control studies or retrospective cohort studies.
- **Level 4 (Low certainty)**: Case series with or without intervention, or cross-sectional studies.
- **Level 5 (Very low certainty)**: Opinion of respected authorities, narrative reviews, case reports, or textbook knowledge.

#### Readability rating

To support public understanding, a medical writer assessed readability and refined simplified statements to a high school reading level and key stakeholder groups, without altering the original scientific statements.

#### Reproducibility

The data supporting the findings of this study, including anonymized voting outcomes and finalized consensus statements, are available at https://github.com/tscnlab/SpitschanEtal_BMJPublicHealth_2025^20^. The data are licensed under the CC-BY 4.0 License, and the code is licensed under the MIT License.

## Results

### Initial statement development

The 27 initial statements aimed to deliver clear, evidence-informed messages about the non-image-forming effects of light. They address a wide range of topics, including the properties of light, how the human eye detects and processes light, and the influence of light on various physiological and psychological functions, such as the regulation of circadian rhythms, sleep, mood, and alertness. The statements also consider how these responses may change with age and include reflections on the current state of research in the field.

A consensus threshold of over 75% agreement was met for all but two of the original statements:

1. “Light exposure can be described by its intensity: The total amount of energy across all wavelengths from 380 to 780 nm.”
2. “Higher light levels during the daytime reduce the physiological effects of light in the evening and at night.”

These statements did not meet the predefined consensus threshold, indicating variability in expert agreement regarding their formulation or interpretation (**Figure 2**). The coordinating committee carefully reviewed all contextual feedback provided by the Consortium. Open-ended feedback revealed that Statement 1 was considered too general, with multiple members recommending the inclusion of information on how the different characteristics of light are weighted. In response, the coordinating committee revised the statement accordingly, and it was subsequently included in the list of agreed-upon statements to be reviewed by the Consortium. In contrast, Statement 2 was deemed to lack sufficient scientific evidence and was therefore removed from the remainder of the consensus process.

**Figure 2.**
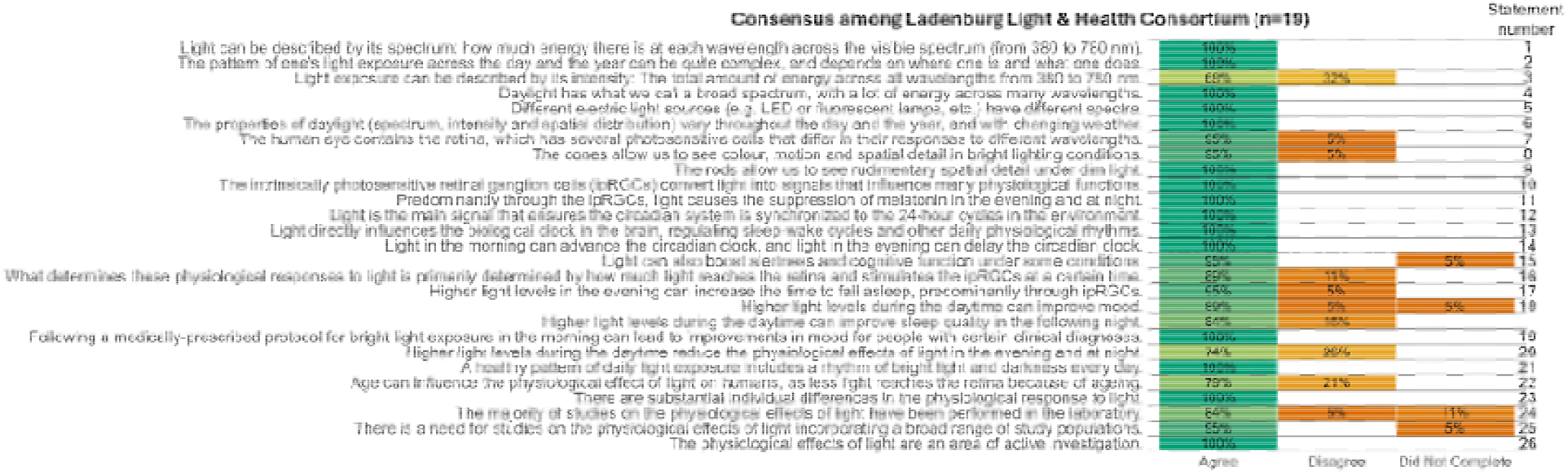
Consensus process and expert agreement levels. This figure illustrates the consensus process for 27 statements on the non-image-forming effects of light, displaying the proportion of participants (n=19) who agreed (green), disagreed (orange), or did not provide a response (orange). Horizontal bars display agreement levels for each statement, with darker shades indicating higher percentages. The consensus threshold is set at 75%. Statements that did not meet this threshold were either amended following contextual feedback or excluded from further consideration. The figure highlights areas of strong agreement as well as topics where expert opinions varied, reflecting the complexity of certain concepts in light and health research.

Additionally, three of the original statements remained unrated by two participants, leading to missing data in the final assessment. However, even if these unrated items were categorized as “disagreed”, the affected statements would still not fall below the 75% consensus criterion. Therefore, they were retained the final set of statements (**Table 1** and **Supplementary Table 1** with simplified statements and contextual information, **Figure 2**).

**Table 1:**
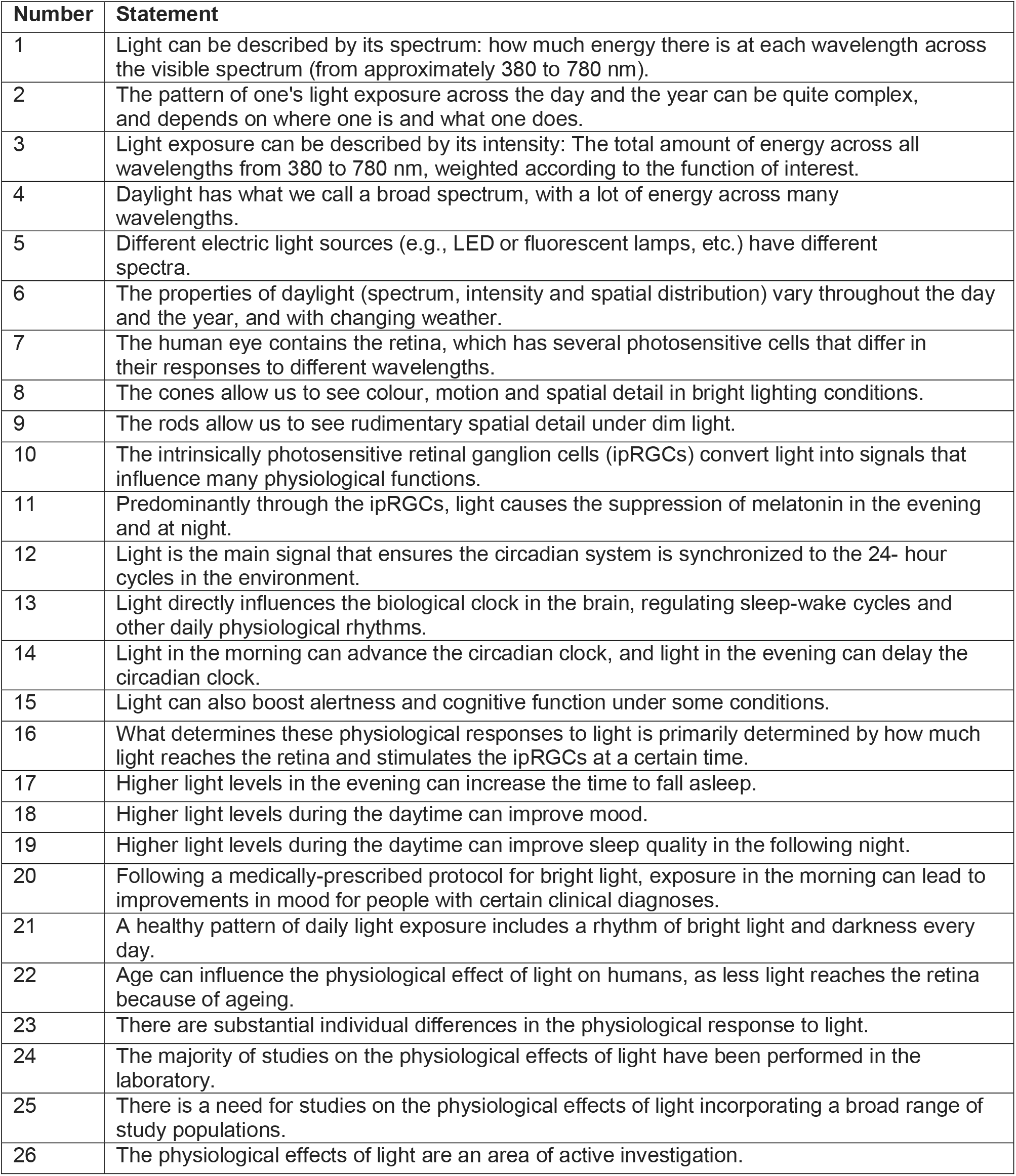
Public health messages on the non-visual effects of ocular light exposure. The “Statement” column provides detailed descriptions using technical terminology. SImplified statements and contextual information is given in **Supplementary Table 1**.

### Consensus statements for public dissemination

The consensus process resulted in the development of 26 statements that were unanimously agreed upon, focusing on the effects of light on humans. These statements provide scientifically supported insights into how light influences physiological and psychological processes in addition to vision. Additionally, the consensus process involved refining these statements into simplified versions to enhance accessibility for a broader audience. Contextual information, along with relevant references, was also incorporated to provide background knowledge and real-world applications, ensuring a comprehensive understanding of the effects of light.

### Evidence rating

The quality of the supporting scientific literature was assessed using a scoring system adapted from the Oxford Centre for Evidence-Based Medicine (OCEBM) framework for rating individual studies. This modified approach allowed for consistent evaluation of the strength and reliability of the evidence cited supporting each statement, considering study design, methodological rigour, and relevance to the topic **(Supplementary Table 2)**. Several statements were supported primarily by textbook knowledge or widely accepted biological principles, where empirical clinical trials are not feasible. This reflects a limitation in the research method rather than a lack of confidence.

### Readability of statements

All scientific statements, simplified statements, and contextual information were assessed for readability by an independent medical writer to ensure accessibility for a broad, non-specialist audience. The Flesch-Kincaid Grade Level for the statements was 8.6, indicating an approximate 8th-grade reading level, with a Flesch Reading Ease score of 59.3, suggesting the content is moderately easy to understand for the general public. On average, the statements contained 14.8 words per sentence and 5 characters per word. The contextual information had a Flesch-Kincaid Grade Level of 10.3 and a Reading Ease score of 43.8, reflecting a slightly more complex reading level. These sections averaged 13 words per sentence and 5.5 characters per word. In contrast, the full scientific versions of the statements were written at a Flesch-Kincaid Grade Level of 11.6, with a Reading Ease score of 42.5, indicating more advanced readability suitable for a specialist audience.

## Discussion

Effective communication of scientific knowledge to the public is essential, especially when it involves behavioural changes with significant implications for health and well-being. Translating complex scientific findings about the effects of light on health into accessible information is critical for promoting awareness and informed decision-making. This public education initiative led to the creation of 26 consensus-based statements on the effects of light on humans, explicitly designed for public dissemination. A high level of agreement (>75%) was achieved for most statements among a group of scientists from different disciplines, with only two exceptions, highlighting areas requiring further discussion and clarification. The iterative process of revising the scientific statements, simplified statements, and contextual information, in collaboration with co-authors, the consortium, and a medical writer skilled in communicating with lay audiences, helped refine the language to improve accessibility and understanding for diverse readers. This collaborative effort resulted in a measurable improvement in readability scores, ensuring that the final materials were not only scientifically accurate but also suitable for effective dissemination to the general public, educators, and policymakers. The improvement in reading level reflects a critical step toward bridging the gap between complex scientific evidence and real-world application.

### Strengths and limitations

One of the key strengths of this initiative is that it represents the first set of consensus-based statements specifically designed for public engagement on light and health. The expert-driven modified Delphi process included input from multiple disciplines, ensuring that a broad range of perspectives was incorporated. The statements underwent an iterative review and refinement process, enhancing their accuracy and clarity. Additionally, readability testing was integrated to ensure accessibility across varying levels of literacy and scientific familiarity. Collaborations with international organizations further strengthened the potential for broad dissemination and impact, increasing the likelihood that the findings will reach and influence a wide audience. A potential limitation of this work is the presence of selection bias, as the roundtable participants represented specific disciplinary backgrounds and may not fully capture the breadth of perspectives. Participation from medical doctors was limited, which may have reduced the clinical input into the health-related recommendations. In addition, experts from low- and middle-income countries (LMICs) were underrepresented, potentially limiting the global applicability of the consensus statements. For statements 18–21, it is especially important to consider regions where high levels of ultraviolet (UV) radiation pose additional risks; in such settings, recommendations for increased light exposure must be carefully balanced with sun-protection guidance. Future initiatives could strengthen the generalizability of the statements by engaging a more diverse range of stakeholders, including greater representation from clinical experts and regional voices. Additionally, there was a necessary trade-off between scientific precision and accessibility, leading to some degree of simplification in the messaging. Further validation of the statements is needed, including public feedback and empirical testing to assess their effectiveness in real-world communication settings. Another limitation is that the process did not include a lay audience in an iterative feedback loop. Incorporating direct input from the general public can provide valuable insights into how non-experts interpret and understand the statements, improving their clarity and impact.

Assessing the quality of evidence on the effects of light on human physiology and psychology presents unique challenges. Much of the foundational work in this field comes from laboratory studies, observational research, and narrative reviews, which may not meet the highest levels of evidence grading systems such as the OCEBM Levels of Evidence we used here. Moreover, the field’s interdisciplinary nature, spanning chronobiology, psychology, lighting engineering, and public health, means diverse study designs and outcome measures are used, complicating harmonized assessment. While formal grading frameworks offer a structure for evaluation, they can undervalue high-quality mechanistic, observational, or exploratory research. While RCTs are often regarded as the highest tier of clinical evidence, in the field of light and health, they are not always feasible, ethical, or appropriate. In such cases, non-RCT designs provide indispensable insights, and lower levels in the OCEBM hierarchy do not necessarily imply weaker relevance or reliability. Additionally, several of the statements developed here represent textbook knowledge, for which a clinical trial or similar (bio)medical study designs are unsuitable. Our use of the OCEBM framework was intended to promote transparency in evidence appraisal, but the interdisciplinary nature of this field requires integrating diverse forms of evidence to inform public health messaging.

### Importance of public engagement in light and health

The urgency of effectively communicating the effects of light on health stems from significant changes in the modern light environment. Increased electric lighting^21^, prolonged screen exposure^22^, and a growing disconnect from natural light-dark cycles have altered human exposure patterns to light and darkness^23^.

Uncertainty persists regarding several aspects of the role of light in circadian health, sleep, and overall well-being. For instance, the consortium did not reach consensus on a statement addressing the extent to which daytime light exposure may counteract the effects of light at night, a topic that is only beginning to be explored in scientific research^24,25^.

Clear, evidence-based messaging on the health impacts of light can empower individuals to make informed choices while also guiding policymakers in shaping workplace environments, urban design, and lighting standards. Through well-structured communication, individuals and decision-makers can be encouraged to prioritize light exposure in ways that support circadian health and overall well-being.

### Plans for implementation

Building on the consensus statements and international collaborations, the next steps involve distributing the statements through organizational networks. A key milestone was the initiative’s official launch on the UNESCO International Day of Light (16 May 2025). Further actions include(1) developing accessible outreach materials such as infographics, social media campaigns, and policy briefs to support dissemination in cooperation with public-health multipliers^17^; (2) translating statements into multiple languages to maximize accessibility and reach diverse populations; (3) establishing predefined indicators to monitor engagement and assess the impact of the communication efforts. Evaluation will include metrics such as website traffic, downloads, and social media analytics; surveys or focus groups to assess comprehension and uptake among target audiences; and tracking references or incorporation of the statements into institutional policies, guidelines, or professional recommendations. Together, these measures will enable assessment of both reach and real-world influence of the initiative. Future iterations will also involve direct testing with members of the public to optimize the clarity, accessibility, and effectiveness of the communication materials.

## Conclusion

This expert consensus delivers clear and accessible messages on the impact of light on human health. By translating scientific evidence into practical guidance, it serves as a valuable resource for public education and policy development. Public health stakeholders, including schools, employers, healthcare providers, and urban planners, can use these messages to support healthier light exposure in everyday life. The statements emphasize the need to consider light as a crucial factor in health, alongside sleep, nutrition, and physical activity.

## Supporting information

Response to reviewers

## Data Availability

All data produced are available online at https://github.com/tscnlab/SpitschanEtal_BMJPublicHealth_2025

https://lightforpublichealh.org/

## Funding

The Ladenburg Roundtable “Light for health and well-being – from the biological principles to policy” (14-16 April 2024) was supported financially by the Daimler and Benz Foundation. The funders had no role in the content, decision to publish, or preparation of the statements. This work was also non-financially supported by International Commission on Illumination (Commission Internationale de l’Éclairage; CIE), the Society for Light, Rhythms and Circadian Health (SLRCH), the Daylight Academy (DLA), the Good Light Group (GLG), and the Center for Environmental Therapeutics (CET).

## Data sharing statement

The data supporting the findings of this study, including anonymized voting outcomes and finalized consensus statements, are available at https://github.com/tscnlab/SpitschanEtal_BMJPublicHealth_2025. The data are licensed under the CC-BY 4.0 License, and the code is licensed under the MIT License. All materials used to develop public-facing statements, including simplified text and contextual explanations, are included in this manuscript and public dissemination materials. No individual-level or sensitive data were collected or analyzed as part of this consensus process.

## Author contributions statement

Manuel Spitschan contributed to the conceptualization, methodology, investigation, writing of the original draft, review and editing, as well as project administration. Laura Kervezee, Oliver Stefani, and Jennifer A. Veitch each contributed to conceptualization, methodology, investigation, and the writing of both the original draft and subsequent review and editing. Marijke Gordijn contributed to conceptualization, methodology, investigation, writing of the original draft, review and editing, and also supported project administration. Renske Lok contributed to conceptualization, methodology, investigation, visualization, and the writing of the original draft, as well as review and editing. The Light for Public Health Consortium contributed to writing, review, and editing.

## Guarantorship

M.S. accepts full responsibility for the finished work and/or the conduct of the study, had access to the data, and controlled the decision to publish.

## Conflict of interest statements

**M.S**. declares the following potential conflicts of interest in the past five years (2021–2025). **Academic roles**: Member of the Board of Directors, *Society of Light, Rhythms, and Circadian Health (SLRCH)*; Chair of *Joint Technical Committee 20 (JTC20)* of the *International Commission on Illumination (CIE)*; Member of the *Daylight Academy*; Chair of *Research Data Alliance Working Group Optical Radiation and Visual Experience Data*. **Remunerated roles**: Speaker of the Steering Committee of the *Daylight Academy*; Ad-hoc reviewer for the *Health and Digital Executive Agency* of the *European Commission*; Ad-hoc reviewer for the *Swedish Research Council*; Associate Editor for *LEUKOS*, journal of the *Illuminating Engineering Society*; Examiner, *University of Manchester*; Examiner, *Flinders University*; Examiner, *University of Southern Norway*. **Funding**: Received research funding and support from the *Max Planck Society, Max Planck Foundation, Max Planck Innovation, Technical University of Munich, Wellcome Trust, National Research Foundation Singapore, European Partnership on Metrology, VELUX Foundation, Bayerisch-Tschechische Hochschulagentur (BTHA), BayFrance (Bayerisch-Französisches Hochschulzentrum), BayFOR (Bayerische Forschungsallianz)*, and *Reality Labs Research*. **Honoraria for talks**: Received honoraria from the *ISGlobal, Research Foundation of the City University of New York* and the *Stadt Ebersberg, Museum Wald und Umwelt*. **Patents**: Named on European Patent Application EP23159999.4A (“*System and method for corneal-plane physiologically-relevant light logging with an application to personalized light interventions related to health and well-being*”). M.S. declares no influence of the disclosed roles or relationships on the work presented herein.

**J.A.V**. declares the following potential conflicts of interest in the past three years (2023–2025). **Academic roles**: President of the *International Commission on Illumination (CIE)*; this is an unpaid role in a global non-profit scientific and standardization organization in the field of light and lighting. J.A.V. declares no influence of the disclosed roles or relationships on the work presented herein.

**L.K**. declares no potential conflicts of interest in the past three years (2023–2025).

**M.G**. declares the following potential conflicts of interest in the past three years (2023–2025). **Employment**: *Chrono@Work B*.*V*. (salary received as an employee). **Academic roles**: Member of the *Good Light Group* (unpaid). M.G. declares no influence of the disclosed roles or relationships on the work presented herein.

**O.S**. declares the following potential conflicts of interest in the past five years (2023–2025). T**ravel support**: Reimbursement for attending the Ladenburg Roundtable “Light for health and well-being – from the biological principles to policy” (April 2025), supported by the *Daimler and Benz Foundation*. **Academic roles**: Member of the Board of Directors, *Schweizer Licht Gesellschaft (Swiss Lighting Society)*. O.S. declares no influence of the disclosed roles or relationships on the work presented herein.

**R.L**. declares the following potential conflicts of interest in the past five years (2023–2025). **Funding**: Salary supported by the *National Institute on Aging (NIA)* through the NIH Pathway to Independence Award (K99/R00), grant number K99AG08484. **Travel support**: Financial support received from the *Center for Environmental Therapeutics* (for SLTBR 2024, Prague) and from *Sleep Europe Congress* (2024, Sevilla). **Academic roles**: Board Member (2018– 2024) and Vice President (from 2024), *Society for Light, Rhythms, and Circadian Health*. R.L. declares no influence of the disclosed roles or relationships on the work presented herein.

## Notes

### Summary of Updates

Included Figures 1 and 2, as well as supplementary files.

