## Supplementary Table S1 for "Evidence-based public health messaging on the non-visual effects of ocular light exposure: A modified Delphi expert consensus"

**Supplementary Table 1: Public health messages on the non-visual effects of ocular light exposure.** The "Statement" column provides detailed descriptions using technical terminology. The "Simplified statement" column translates these concepts into more accessible language. The "Contextual information" column provides additional background and real-world examples with references to enhance comprehension and illustrate the relevance of the concepts.

| **Number** | **Statement** | **Simplified statement** | **Contextual information** |
| --- | --- | --- | --- |
| 1 | Light can be described by its spectrum: how much energy there is at each wavelength across the visible spectrum (from approximately 380 to 780 nm). | White light is made up of multiple wavelengths, which we perceive as colours. | When we see different colours, we are actually seeing different wavelengths of light. Short wavelengths appear violet, while longer wavelengths appear indigo, blue, green, yellow, orange, and red. |
| 2 | The pattern of one's light exposure across the day and the year can be quite complex,  and depends on where one is and what one does. | The amount of light around us (or the amount we are exposed to) changes as we move between indoor and outdoor spaces, throughout the day, and across seasons. | Light from the sun increases from dawn to midday and decreases in the evening. Depending on location, the amount of light in the day varies greatly between summer and winter. |
| 3 | Light exposure can be described by its intensity: The total amount of energy across all  wavelengths from 380 to 780 nm, weighted according to the function of interest. | We can measure light exposure by assessing how intense the light is at different wavelengths. | We can characterise a light source by describing how much power it emits at each wavelength. This power is called intensity. Physicists measure intensity, while biological and psychological scientists study how intensity affects various processes. |
| 4 | Daylight has what we call a broad spectrum, with a lot of energy across many  wavelengths. | Daylight has a wide range of energy across multiple wavelengths – it can be separated into all the colours of the rainbow. | Daylight, a combination of direct sunlight and scattered sky light, contains all visible wavelengths and more. Changes in the time of day and weather conditions shift the wavelength composition. This shift changes the colour that daylight appears. |
| 5 | Different electric light sources (e.g., LED or fluorescent lamps, etc.) have different  spectra. | Different light sources generate light in different ways, producing distinct wavelengths or colour. | Different light source technologies use various materials to convert electricity into light. Different materials cause the intensity of light to spread across different wavelengths in different ways. This spread of light across wavelengths is known as spectral distribution. |
| 6 | The properties of daylight (spectrum, intensity and spatial distribution) vary throughout the day and the year, and with changing weather. | The colour, intensity, and pattern of daylight change throughout the day, with seasons, and weather. | As sunlight travels through the atmosphere, blue light is scattered more than other wavelengths. This makes the sky appear blue. |
| 7 | The human eye contains the retina, which has several photosensitive cells that differ in  their responses to different wavelengths. | In the eye, the retina contains cells that allow us to detect different colours. | In the human eye, the retina contains light-sensitive cells that convert light into signals for the brain. These cells are called cones, rods, and intrinsically photosensitive retinal ganglion cells (ipRGCs)^26,27^. |
| 8 | The cones allow us to see colour, motion and spatial detail in bright lighting conditions. | Cone cells help us see colours, movement, and objects in bright light. | Cones are specialized cells in the retina. They are named for their shape (how they appear under a microscope). The highest density of cones is in the fovea (in the centre of the retina). |
| 9 | The rods allow us to see rudimentary spatial detail under dim light. | Rod cells help us see shapes and details in dim light. | Rods are highly sensitive to low light levels. They are essential for seeing at night. |
| 10 | The intrinsically photosensitive retinal ganglion cells (ipRGCs) convert light into signals that influence many physiological functions. | When ipRGCs detect light, they send signals to the brain to regulate various bodily functions. | ipRGCs send electrical signals to brain regions that regulate sleep-wake cycles, alertness, and mood. |
| 11 | Predominantly through the ipRGCs, light causes the suppression of melatonin in the evening and at night. | Light blocks the production of melatonin (a hormone that regulates sleep-wake cycles), particularly in the evening and at night. | ipRGCs express melanopsin (a light-sensitive protein). When ipRGCs detect bright light, melanopsin becomes activated. This activation triggers a neuronal pathway, blocking the production of melatonin in the pineal gland. |
| 12 | Light is the main signal that ensures the circadian system is synchronized to the 24- hour cycles in the environment. | Light is the primary signal that synchronizes the body's internal clock with the sun’s day-night cycle. | Light helps regulate the body's internal rhythms. This ensures biological processes start and stop at appropriate times. |
| 13 | Light directly influences the biological clock in the brain, regulating sleep-wake cycles and other daily physiological rhythms. | Light influences the body’s internal clock, which regulates sleep-wake patterns and other daily rhythms. | Light exposure influences sleep, hormone release, metabolism, and alertness. |
| 14 | Light in the morning can advance the circadian clock, and light in the evening can delay the circadian clock. | Morning light promotes earlier bedtime and wake time, while evening light can delay bedtime and waking. | Light tells the brain to become active. Light in the morning advances the body’s internal clock (shifts it to an earlier “time”), while light in the evening delays it (shifts it to a later “time”). |
| 15 | Light can also boost alertness and cognitive function under some conditions. | In certain situations, light can improve alertness and ability to think. | Under some conditions, exposure to high-intensity light during the day and night enhances alertness and improves cognitive performance. |
| 16 | What determines these physiological responses to light is primarily determined by how much light reaches the retina and stimulates the ipRGCs at a certain time. | The body's response to light depends on how much light and what time the light reaches the ipRGCs in the retina. | The response of ipRGCs likely depends on the time of day and circadian factors. |
| 17 | Higher light levels in the evening can increase the time to fall asleep. | Evening light exposure can make it harder to fall asleep. | Evening light exposure tells the body it is still daytime. This increases alertness and shifts the internal clock to a later time. |
| 18 | Higher light levels during the daytime can improve mood. | Bright light during the day can improve mood. | Exposure to natural daylight (the sun) or high-intensity electric light, when free from glare, can reduce stress and improve emotional balance. |
| 19 | Higher light levels during the daytime can improve sleep quality in the following night. | Exposure to more intense light during the day can improve sleep. | Exposure to higher daytime light levels reduces sleep fragmentation (awakening throughout the night) and increases deep sleep at night. |
| 20 | Following a medically-prescribed protocol for bright light, exposure in the morning can lead to improvements in mood for people with certain clinical diagnoses. | Doctors may prescribe light therapy as treatment for winter depression and other health conditions. | Exposure to bright light has been shown to improve depressive symptoms in individuals with Seasonal Affective Disorder. |
| 21 | A healthy pattern of daily light exposure includes a rhythm of bright light and darkness every day. | A healthy daily routine includes bright light during the day and darkness at night. | Maintaining a regular pattern of bright light during the day and darkness at night is associated with better physical and mental health. |
| 22 | Age can influence the physiological effect of light on humans, as less light reaches the retina because of ageing. | As people age, the lenses of the eye can become less transparent, which may reduce the effects of light on the internal clock and sleep. | With age, the eye’s lenses become less transparent, which may reduce the body’s response to light exposure. |
| 23 | There are substantial individual differences in the physiological response to light. | How people respond to light can vary greatly. | Individual differences in light sensitivity are influenced by factors such as age, genetics, and behavior. |
| 24 | The majority of studies on the physiological effects of light have been performed in the laboratory. | Most research on light’s effects on the body has been conducted in the laboratory. | Studies investigating light exposure in the real-world are needed. |
| 25 | There is a need for studies on the physiological effects of light incorporating a broad range of study populations. | More studies are needed to understand how light influences health in different groups of people. | Most studies on the physiological effects of light have focused on limited populations (specific age groups, ethnicities, or health conditions). It is important that future research studies more diverse populations over wider geographical regions. |
| 26 | The physiological effects of light are an area of active investigation. | Scientists continue to explore how light affects bodily functions and overall health. | Scientific and popular interest in the effects of light on biological rhythms, sleep, alertness, mood, and health is growing. Technological and scientific advances are enabling researchers to more precisely study different wavelengths and different intensities. |
