## Supplementary Table S2 for "Evidence-based public health messaging on the non-visual effects of ocular light exposure: A modified Delphi expert consensus"

**Supplementary Table 2: Rating of the quality of evidence.** This table lists each reference, corresponding number, general study type, and an evidence quality rating assigned by two independent raters (authors ML and RL). Ratings are based on a modified version of the Oxford Centre for Evidence-Based Medicine levels for individual studies. The five levels of evidence are defined as follows: (1) Properly powered and conducted randomized clinical trial or systematic review with meta-analysis; (2) Well-designed controlled trial without randomization or prospective comparative cohort trial; (3) Case-control studies or retrospective cohort study; (4) Case series with or without intervention or cross-sectional study; (5) Opinion of respected authorities or case reports. As many of the statements represent textbook knowledge of basic physics and biology, we have added the rating category (–) for these statements.

| **Number** | **Statement** | **Main supporting sources** | **Rating** |
| --- | --- | --- | --- |
| 1 | Light can be described by its spectrum: how much energy there is at each wavelength across the visible spectrum (from approximately 380 to 780 nm). | ^28,29^ | – |
| 2 | The pattern of one's light exposure across the day and the year can be quite complex, and depends on where one is and what one does. | ^30^ | – |
| 3 | Light exposure can be described by its intensity: The total amount of energy across all wavelengths from 380 to 780 nm, weighted according to the function of interest. | ^31^ | – |
| 4 | Daylight has what we call a broad spectrum, with a lot of energy across many wavelengths. | ^29^ | – |
| 5 | Different electric light sources (e.g., LED or fluorescent lamps, etc.) have different spectra. | ^32,33^ | – |
| 6 | The properties of daylight (spectrum, intensity and spatial distribution) vary throughout the day and the year, and with changing weather. | ^33-36^ | – |
| 7 | The human eye contains the retina, which has several photosensitive cells that differ in their responses to different wavelengths. |  | – |
| 8 | The cones allow us to see colour, motion and spatial detail in bright lighting conditions. | ^37,38^ | – |
| 9 | The rods allow us to see rudimentary spatial detail under dim light. | ^39,40^ | – |
| 10 | The intrinsically photosensitive retinal ganglion cells (ipRGCs) convert light into signals that influence many physiological functions. | ^26,41,42^ | – |
| 11 | Predominantly through the ipRGCs, light causes the suppression of melatonin in the evening and at night. | ^43-45^ | 1 |
| 12 | Light is the main signal that ensures the circadian system is synchronized to the 24-hour cycles in the environment. | ^2,4,22,46^ | 2 |
| 13 | Light directly influences the biological clock in the brain, regulating sleep-wake cycles and other daily physiological rhythms. | ^2,4,22^ | 2 |
| 14 | Light in the morning can advance the circadian clock, and light in the evening can delay the circadian clock. | ^6,7^ | 2 |
| 15 | Light can also boost alertness and cognitive function under some conditions. | ^8,9,12^ | 2 |
| 16 | What determines these physiological responses to light is primarily determined by how much light reaches the retina and stimulates the ipRGCs at a certain time. | ^45,47-50^ | 2 |
| 17 | Higher light levels in the evening can increase the time to fall asleep. | ^50-54^ | 1 |
| 18 | Higher light levels during the daytime can improve mood. | ^23,55^ | 3 |
| 19 | Higher light levels during the daytime can improve sleep quality in the following night. | ^56-58^ | 2 |
| 20 | Following a medically-prescribed protocol for bright light, exposure in the morning can lead to improvements in mood for people with certain clinical diagnoses. | ^59,60^ | 1 |
| 21 | A healthy pattern of daily light exposure includes a rhythm of bright light and darkness every day. | ^15,24,61,62^ | 3 |
| 22 | Age can influence the physiological effect of light on humans, as less light reaches the retina because of ageing. | ^63-66^ | 3 |
| 23 | There are substantial individual differences in the physiological response to light. | ^54,67-69^ | 2 |
| 24 | The majority of studies on the physiological effects of light have been performed in the laboratory. | ^70^ | 5 |
| 25 | There is a need for studies on the physiological effects of light incorporating a broad range of study populations. | ^71,72^ | 5 |
| 26 | The physiological effects of light are an area of active investigation. | ^73^ | 5 |
