## Supplementary material for "Evidence-based public health messaging on the non-visual effects of ocular light exposure: A modified Delphi expert consensus": Response to reviewers

**Spitschan et al., BMJ Public Health**

Response to reviewers

**Editor comments**

We thank the editor and reviewers for their constructive comments on our manuscript. We have revised the manuscript following the comments, and list the changes below. We feel that the manuscript has substantially improved after these comments.

In addition to the changes made, we also note that the consortium name has changed to "Light for Public Health Consortium", which we have updated in the manuscript for accuracy.

| **Comment** | **Comment** | **Response** |
| --- | --- | --- |
| E1.1 | Please include a statement relating to the ethical approval obtained for your study. If ethical approval was not required, please explain why this is the case. Please also provide a statement regarding participant consent. | An ethics statement was added to the manuscript, page 4:  *Ethics. This research involved expert participants contributing in their professional capacity to a consensus process. No clinical interventions, personal health data, or sensitive information were collected. Formal ethical approval was therefore not required under prevailing guidelines. All participants were informed about the purpose and procedures of the study and provided consent through their voluntary agreement to take part.* |
| E1.2 | Please ensure that your abstract is formatted according to our Original Research submission guidelines: https://bmjpublichealth.bmj.com/pages/authors#original_research | The abstract has been reformatted in accordance with BMJ Public Health guidelines. |
| E1.3 | Inspired by the work of the patient partnership strategy at The BMJ (https://www.bmj.com/campaign/patient-partnership), BMJ Public Health is encouraging active patient involvement in setting the research agenda. BMJ Public Health requires authors of all submissions to the journal to include a Patient and Public Involvement statement. The Patient and Public Involvement statement should be included as a sub-heading in the methods section of all manuscripts. It should provide a brief description of any patient involvement in study design or conduct of the study, as well as any plans to disseminate the results to study participants. If patients and or the public were not involved, please state "None". See the our Instructions for Authors for further details: https://bmjpublichealth.bmj.com/pages/authors#reporting_patient | The following has been added:  *Patient and Public Involvement statement*  *Patients or members of the public were not involved in the design, conduct, reporting, or dissemination of this research. The consensus process was restricted to expert participants selected for their scientific expertise in light, health, and related disciplines. An independent medical writer contributed to refining the public-facing statements to ensure accessibility and clarity.* |
| E1.4 | Please provide more details in the methods regarding the rating procedure and scale. | We have now added methodological details in the methods section. |
| E1.5 | Key message Please include the key messages of your article after your abstract using the following headings. This section should be no more than 3-5 sentences and should be distinct from the abstract; be succinct, specific and accurate.  What is already known on this topic – summarise the state of scientific knowledge on thics subject before you did your study and why this study needed to be done   What this study adds – summarise what we now know as a result of this study that we did not know before  How this study might affect research, practice or policy – summarise the implications of this study | We have now added the following statements:  *Light exposure has an impact on human health beyond vision, affecting sleep, alertness and circadian rhythms. However, policy guidance on healthy lighting is limited.*  *This expert consensus document sets out 26 key messages on the health impacts of light in clear, accessible language.*  *These messages provide policymakers with reliable guidance to help create healthy lighting environments. They can inform regulations, public awareness campaigns and health promotion strategies.* |

| **Comment** | **Reviewer comment** | **Response** |
| --- | --- | --- |
| R1.1 | This paper addresses an important and timely public health issue. With the increasing use of smartphones, smartwatches, and other digital devices, not only for communication but also for physiological monitoring, entertainment, and work, exposure to artificial light has increased significantly. This prolonged exposure to light of varying wavelengths and intensities, particularly outside of natural circadian patterns, has the potential to impact sleep, mood, and long-term health. Yet, public awareness remains limited. Thus, the effort to formulate accessible, evidence-informed public health statements on this topic is needed.  The manuscript is clearly structured, and the consensus-building process is described in detail. The iterative development of scientifically accurate yet simplified public messages is a strength. However, I would like to highlight several points for consideration and improvement: | We thank the reviewer for recognizing the importance of this topic and the strengths of our consensus-building approach, and we appreciate the constructive suggestions for further improvement |
| R1.2 | *Representation of Expertise and Geography* While the Delphi panel includes a multidisciplinary group of scientists, it lacks participation from medical doctors, which is notable given the health-oriented focus of the recommendations. Moreover, there is limited representation from low- and middle-income countries, which may limit the generalizability and practical applicability of the consensus statements across diverse healthcare systems and populations - particularly given the authors’ plans for global outreach through organizations such as UNESCO. | We have added the following statement to the discussion:  *A potential limitation of this work is the presence of selection bias, as the roundtable participants represented specific disciplinary backgrounds and may not fully capture the breadth of perspectives. In particular, participation from medical doctors was limited, which may have reduced the clinical input into the health-related recommendations. In addition, experts from low- and middle-income countries (LMICs) were underrepresented, potentially limiting the global applicability of the consensus statements. For statements 18–21, it is especially important to consider regions where high levels of ultraviolet (UV) radiation pose additional risks; in such settings, recommendations for increased light exposure must be carefully balanced with sun-protection guidance. Future initiatives could strengthen the generalizability of the statements by engaging a more diverse range of stakeholders, including greater representation from clinical experts and regional voices.* |
| R1.3 | *Lack of Systematic Review* A major limitation is that no systematic literature review was conducted to identify and evaluate the evidence used as a basis for the statements. While the modified Delphi method is valid for expert consensus, the absence of a systematic search process introduces a risk of selection bias and this too should be clearly acknowledged in the limitations section of the paper and the alternative method used to review the relevant literature should be described in the materials and methods section. | We appreciate the reviewer’s comment and agree that systematic reviews are valuable in many contexts; however, in this case a systematic review was neither feasible nor appropriate, as the evidence base for light and health is highly interdisciplinary, heterogeneous in study designs, and includes foundational physiological principles and standards that cannot be captured through conventional systematic review approaches. Instead, we employed a structured, iterative consensus process drawing on the expertise of an international, multidisciplinary panel, which is an established and rigorous method for synthesizing evidence and formulating consensus-based recommendations in areas where the literature is broad, complex, and not easily amenable to systematic synthesis. We have clarified this rationale in the Methods and noted it as a limitation, while emphasizing the complementary role of future systematic reviews to address more focused research questions. |
| R1.4 | *Transparency of Evidence Ratings Method* While a modified version of the Oxford Centre for Evidence-Based Medicine (OCEBM) framework was used to rate evidence quality, it is not clear whether this modification has been validated or previously published. Greater transparency is needed regarding:   - How the modifications differ from the original OCEBM system. - Who performed the evidence ratings and under what criteria. - How evidence ratings are communicated on the associated website (lightforpublichealth.org), where certainty levels are currently not visible. | We thank the reviewer for this comment and would like to clarify that we relied on the 2011 update of the Oxford Centre for Evidence-Based Medicine (OCEBM) Levels of Evidence, which itself represents a modified version of the original 1998 framework. The other update is that we added (-) for textbooks.  Authors RL and MS provided the ratings, which has been added to the manuscript: *This table lists each reference, corresponding number, general study type, and an evidence quality rating assigned by two independent raters (ML and RL).* The criteria are outlined in the table legend.  Upon acceptance of this manuscript as a peer-reviewed publication, we will amend the website to include the ratings. |
| R1.5 | *Risk of Confirmation Bias* The coordinating committee developed the initial statements, led revisions, and represented endorsing institutions. This centralized control, while efficient, increases the risk of confirmation bias. Inclusion of a more independent or neutral party at the drafting stage could improve methodological objectivity. | We agree that centralized control can introduce risks of confirmation bias. To mitigate this, we employed an independent Delphi process in which the broader group of expert participants who were not part of the coordinating committee provided anonymized ratings and open-text feedback on all draft statements. All revisions were based on this iterative, blinded feedback rather than solely on the views of the coordinating committee. This has been added to the Materials: *To minimize confirmation bias, however, all subsequent revisions were guided by anonymized ratings and open-text feedback from the broader group of expert participants, who were not members of the coordinating committee. This occurred through a two-step Delphi peer-review process.* |
| R1.6 | *Evaluation of Public-Facing Impact* The authors describe plans for dissemination via outreach materials and translations, but do not outline how the real-world impact of these materials will be measured. Information on evaluation indicators, such as public uptake, behavioral changes, or policy influence, would strengthen the translational value of the work. | We have added the following to the discussion: *Evaluation will include metrics such as website traffic, downloads, and social media analytics; surveys or focus groups to assess comprehension and uptake among target audiences; and tracking references or incorporation of the statements into institutional policies, guidelines, or professional recommendations. Together, these measures will enable assessment of both reach and real-world influence of the initiative.* |
| R1.7 | *Readability vs. Comprehension* While the use of Flesch-Kincaid readability scores is a helpful metric, these scores are not a substitute for empirical testing with target audiences. Without formal user testing, it remains uncertain whether the simplified statements truly promote understanding and actionable behavior among laypeople. | We agree with the reviewer that Flesch-Kincaid readability scores cannot substitute for empirical testing; we have now acknowledged this as a limitation of our study and highlight the need for future work involving testing with lay audience. *Further validation of the statements is needed, including public feedback and empirical testing to assess their effectiveness in real-world communication settings.* |
| R1.8 | This paper presents a valuable initiative to bridge the gap between scientific research and public health messaging on light exposure. The consensus statements are likely to be useful to public-health stakeholders, including educators, healthcare providers, and urban planners. However, further attention to inclusivity, evidence transparency, methodological rigor, and public engagement would strengthen the impact and applicability of this work. With these improvements, the initiative could become a benchmark model for other emerging areas of environmental health communication. | We thank the reviewer for this thoughtful summary and for recognizing the value of our initiative. We agree that inclusivity, evidence transparency, methodological rigor, and public engagement are important areas for further strengthening, and we have addressed these points by clarifying our methods, explicitly acknowledging limitations, and outlining future steps to broaden stakeholder involvement and enhance evaluation. We are grateful for the constructive feedback, which we believe has improved the clarity and applicability of the manuscript. |

| **Comment** | **Reviewer comment** | **Response** |
| --- | --- | --- |
| R2.1 | While the OCEBM ratings are used, many statements rely on foundational knowledge or non-RCT designs. | We thank the reviewer for this observation. While we acknowledge that many of the consensus statements are not based on RCTs, this reflects the interdisciplinary and mechanistic nature of research on light and health, where foundational physiological principles, laboratory studies, and observational research provide essential evidence. In this field, RCTs are often impractical or inappropriate (e.g., testing the role of ipRGCs in circadian regulation). Thus, while OCEBM ratings offer transparency, it is important to recognize that lower tiers in the hierarchy do not necessarily imply weaker relevance for public health guidance. We have clarified this point in the Discussion: *Assessing the quality of evidence on the effects of light on human physiology and psychology presents unique challenges. Much of the foundational work in this field comes from laboratory studies, observational research, and narrative reviews, which may not meet the highest levels of evidence grading systems such as the OCEBM Levels of Evidence we used here. Moreover, the field's interdisciplinary nature, spanning chronobiology, psychology, lighting engineering, and public health, means diverse study designs and outcome measures are used, complicating harmonized assessment. While formal grading frameworks offer a structure for evaluation, they can undervalue high-quality mechanistic, observational, or exploratory research. While RCTs are often regarded as the highest tier of clinical evidence, in the field of light and health, they are not always feasible, ethical, or appropriate. In such cases, non-RCT designs provide indispensable insights, and lower levels in the OCEBM hierarchy do not necessarily imply weaker relevance or reliability. Additionally, several of the statements developed here represent textbook knowledge, for which a clinical trial or similar (bio)medical study designs are unsuitable. Our use of the OCEBM framework was intended to promote transparency in evidence appraisal, but the interdisciplinary nature of this field requires integrating diverse forms of evidence to inform public health messaging.* |
| R2.2 | Suggest adding a brief note in the Results or Discussion explicitly stating that although some statements are rated at Level 3 or below, this reflects limitations in research methods rather than a lack of confidence. | This has been added to the section: “Evidence rating” |
| R2.3 | The manuscript acknowledges that no layperson involvement occurred. Suggest adding one line in Plans for Implementation stating that future iterations will involve direct testing with the public to optimize communication effectiveness. | We have added a Patient and Public Involvement statement, and we have added the following to the Plans for Implementation: *Future iterations will also involve direct testing with members of the public to optimize the clarity, accessibility, and effectiveness of the communication materials.”* |
|  | Some simplified statements might still benefit from greater clarity or contextual richness. Examples: Statement 15 ("Light can also boost alertness..."): clarify that this effect is conditional on timing, intensity, and duration. Statement 18 ("Higher light levels...improve mood"): consider citing this recent evidence <https://pubmed.ncbi.nlm.nih.gov/40606287/> in the contextual info. | While initial statements contained that degree of specificity, we had to simplify some of these statements. This was addressed as a limitation: *Additionally, there was a necessary trade-off between scientific precision and accessibility, leading to some degree of simplification in the messaging. Further validation of the statements is needed, including public feedback and empirical testing to assess their effectiveness in real-world communication settings.*  We have checked the citation, which refers to light at night and diabetes risk, which we feel is not relevant for this section. |
| R2.4 | Figure 2 on consensus levels is strong but could be enhanced by briefly noting in the legend which statement numbers correspond to those that failed to meet consensus. | We have now added a separate column indicating the statement number. |
| R2.5 | The reference list is comprehensive, but some non-English or gray literature sources could be added in future iterations to broaden cultural relevance. | We agree that incorporating non-English and gray literature would broaden the cultural relevance of the reference base, and we have noted this as an area for improvement in future iterations of the initiative. |
| R2.6 | Minor Editorial Issues: Typos/Grammar: The manuscript is generally clean. A few phrases could be more concise. For instance:  "Light tells the brain to become active..." might be rephrased to avoid anthropomorphism, e.g., "Light exposure activates alertness-related brain processes." | We have carefully reviewed the manuscript, corrected typographical errors, and streamlined phrasing where appropriate to improve clarity and conciseness.  We recognize the reviewer’s point regarding anthropomorphic phrasing. As the statements represent the agreed-upon consensus wording, we cannot alter them here, but we have noted this for refinement in subsequent consensus rounds. |
| R2.7 | Citation Consistency: Ensure that all supporting statements (especially those in Table 1) have consistent in-text references and match the evidence rating table. | We carefully reviewed the consistency of all references included. |
